# Improved performance of saliva for the detection of the B.1.351 variant in South Africa

**DOI:** 10.1101/2021.03.29.21254563

**Authors:** Chun Yat Chu, Gert Marais, Christoffel Opperman, Deelan Doolabh, Arash Iranzadeh, Carisa Marais, Helen Cox, Carolyn Williamson, Diana Hardie, Adrian Brink

## Abstract

Assessment of the unknown performance of saliva for the detection of the novel SARS-CoV-2 variant of concern (VOC) B.1.351 (501Y.V2) lineage is essential as saliva has been shown to be an equivalent, less invasive and a less costly alternative to nasopharyngeal swabs for the molecular detection of SARS-CoV-2 infection in pre-variant studies. Between 1st August 2020 and 16th January 2021, we enrolled 410 eligible ambulatory participants who presented to Groote Schuur Hospital (GSH) in Cape Town, South Africa for SARS-CoV-2 testing. Of these, 300 were enrolled prior to, and 110 after, the initial detection and replacement of wild-type by the B.1.351 variant. All participants provided a supervised self-collected mid-turbinate (MT) and saliva (SA) swab, in addition to the standard HCW collected NP swab which were all tested by RT-PCR in an accredited diagnostic laboratory. Positive percent agreement to NP swab for SA swabs pre- and post-variant were 51.5% and 72.5% respectively while these values for MT swabs were 75.8% and 77.5%. The negative percent agreement for all swab types during all periods was >98%. The basis for this marked improvement of SA swabs as a diagnostic sample for B.1.351 virus is still being investigated.

## Body

In December 2020, South Africa (SA) faced a surge in COVID-19 cases, which was associated with the replacement of previous circulating lineages with a novel SARS-CoV-2 variant of concern (VOC) B.1.351 (501Y.V2) lineage^1^. Preliminary analyses suggest that this variant, defined by 3 mutations at key sites in the receptor-binding domain, may have functional significance including increased transmissibility^1^. In the assessment of multiple diagnostic sample types to reduce the need for directed healthcare worker (HCW) performed sampling, saliva has been shown to be equivalent, safer and a less costly alternative to nasopharyngeal (NP) swabs for molecular confirmation of SARS-CoV-2 infection^2,3^. Its performance in the detection of VOC virus is, however, unknown.

Between 1^st^ August 2020 and 16^th^ January 2021, we enrolled 410 eligible ambulatory participants who presented to Groote Schuur Hospital (GSH) in Cape Town, South Africa for SARS-CoV-2 testing. Of these, 300 were enrolled prior to, and 110 after, the initial detection and replacement of the B.1.351 variant. All participants provided a supervised self-collected mid-turbinate (MT) and saliva (SA) swab, in addition to the standard HCW collected NP swab which were tested by RT-PCR^4^ in an accredited diagnostic laboratory. Assessment of the diagnostic validity of MT and SA relative to NP swabs (Fig. 1A) was assessed by comparing the mean cycle threshold (Ct) differences (Fig. 1B and C), pre-test probability (Fig 1D). Whole genome sequencing of suitable specimens (Fig. 1E), was performed.

**Figure 1.**
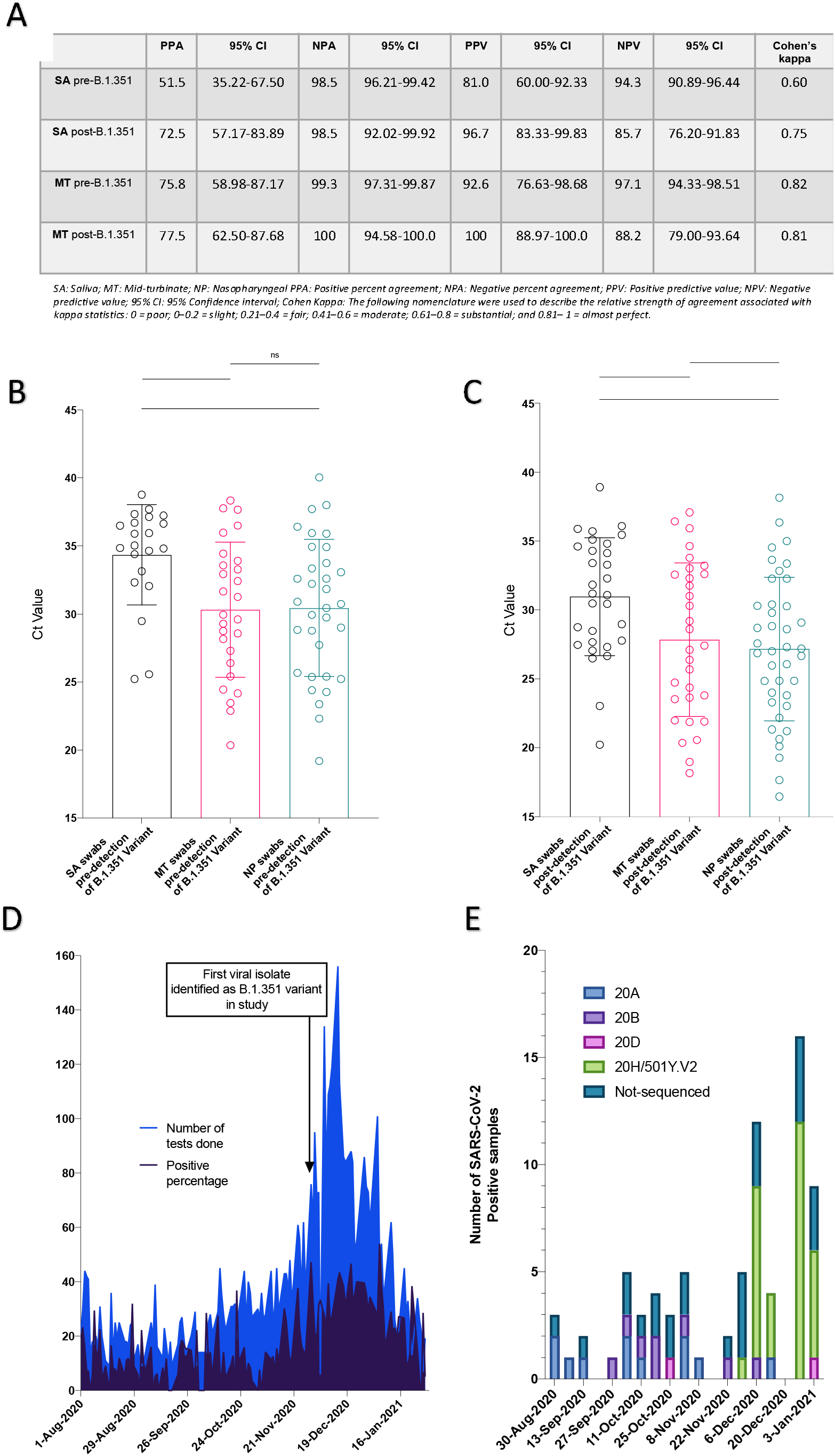
A: Positive and negative percent agreement to nasopharyngeal (NP) swab as well as positive and negative predictive value of saliva (SA) and mid-turbinate (MT) self-administered swabs as a diagnostic sample for SARS-CoV-2 PCR prior to (n=33/300) and after (n=40/107) the emergence of the B.1.351 lineage. Post-emergence, 3 proven non-B.1.351 lineage virus infected participants were excluded. The likelihood ratio of having a positive SA swab result pre- and post-emergence of the B.1.351 lineage for participants with a positive NP swab result was 34 and 48 respectively. Confidence intervals were calculated using the Wilson-Brown method. B and C: Mean cycle threshold (Ct) values for SA, MT and NP swabs are shown with 1 standard deviation error bars. B. Pre-B.1.351 emergence [34.35 (95% CI: 32.67-36.02), 30.31 (95% CI: 28.34-32.27) and 30.44 (95%: 28.65-32.22) respectively] C. Post-emergence [30.96 (95% CI: 29.36-32.56), 27.84 (95% CI: 25.79-29.88), 27.16 (95% CI: 25.50-28.83) respectively]. The mean Ct SA values decreased by 3.39 and a comparable 2.47 and 3.28 for MT and NP, respectively. Paired t-tests were used to compare swab Ct values. *: P-value < 0.05, **: P-value <0.01, ***: P-value <0.001, ****: P-value <0.0001. D: Longitudinal number of all tests performed and positive percentage of SARS-CoV-2 tests over the study period at the GSH testing centre. Participants in this study are included. E: Whole genome sequencing. Longitudinal number of isolates from each clade including the number of viral isolates not deemed suitable for sequencing (Ct values ≥30).

Notably, a noted improvement in the positive percent agreement (PPA) for SA but not MT relative to NP swabs, for the two cohorts was documented. The reasons for the significant change in SA PPA are currently unknown. Possibilities include changes in tissue tropism^5^ associated with the VOC B.1.351, and while increased viral replication in salivary glands would be expected to decrease the mean Ct value to a greater extent than for NP swabs post B.1.351 lineage emergence, this was not evident. However, a potential explanation would be the presence of greater RNA quantities of VOC B.1.351 reflected by decreased mean Ct values for all sample types, which may support the preliminary modelling-based finding of increased transmissibility^1^. Whilst altered test-seeking behavior in the study population cannot be excluded the inclusion criteria remained the same throughout the study.

Whilst further WGS studies are ongoing to determine compartmentalized replication and to inform diagnostic preferences concurrent to public health interventions, knowledge of distinct oral and oropharyngeal shedding dynamics and viral burden through the continuum of SARS-CoV-2 infections is warranted.

## Data Availability

no additional data provided

## Reference

1. Tegally H, Wilkinson E, Giovanetti M, et al. Emergence of a SARS-CoV-2 variant of concern with mutations in spike glycoprotein. Nature 2021.https://doi.org/10.1038/s41586-021-03402-9.

2. Tu Y-P, Jennings R, Hart B, et al. Swabs collected by patients or health care workers for SARS-CoV-2 testing. New England Journal of Medicine 2020;383:494–6.

3. Bastos ML, Perlman-Arrow S, Menzies D, Campbell JR. The Sensitivity and Costs of Testing for SARS-CoV-2 Infection With Saliva Versus Nasopharyngeal Swabs : A Systematic Review and Meta-analysis. Ann Intern Med 2021:M20–6569.

4. Marais G, Naidoo M, Hsiao N-y, Valley-Omar Z, Smuts H, Hardie D. The implementation of a rapid sample preparation method for the detection of SARS-CoV-2 in a diagnostic laboratory in South Africa. PloS one 2020;15:e0241029

5. Huang N, Perez P, Kato T, et al. Integrated single-cell atlases reveal an oral SARS-CoV-2 infection and transmission axis. medRxiv. 27 October, 2020. doi: 10.1101/2020.10.26.20219089. preprint.

